# A prospective case-control study of the gut bacteriome and mycobiome in glioblastoma

**DOI:** 10.64898/2026.09.23.26363766

**Authors:** Michaela Herz, Kim Schmid, Markus Ankenbrand, Elias Roth, Kerstin Hünniger, Nele Gündermann, Robert Nickl, Almuth Friederike Kessler, Andrea Cattaneo, Camelia Maria Monoranu, Ralf-Ingo Ernestus, Carsten Hagemann, Oliver Kurzai, Mario Löhr, Vera Nickl

## Abstract

The gut-brain axis may modulate glioblastoma biology, but human data are scarce and the gut mycobiome remains uncharacterized. In a prospective case-control study, we sequenced bacterial (16S rRNA gene) and fungal (ITS) communities from rectal swabs of 23 patients with newly diagnosed glioblastoma and 23 healthy controls. Fungal diversity was lower in patients than in controls (p < 0.001). Bacterial diversity and overall structure of both communities did not differ. Several short-chain fatty acid producers within the Lachnospiraceae and Ruminococcaceae were nominally reduced in patients, whereas the methanogenic archaeon *Methanobrevibacter* and *Nakaseomyces glabratus* were nominally enriched. In an exploratory multivariate Cox analysis, a higher intratumoral proportion of ionized calcium-binding adapter molecule 1-positive myeloid cells was associated with longer survival, independent of age, performance status and extent of resection (hazard ratio 0.82 per percentage point, p = 0.04). We hypothesize that the myeloid compartment links the changes in gut and tumor.

## Introduction

Glioblastoma (GBM), isocitrate dehydrogenase wildtype, central nervous system (CNS) World Health Organization (WHO) grade 4, is the most frequent malignant primary brain tumor in adults and remains the central unsolved problem of neuro-oncology. It accounts for approximately 52% of primary malignant brain tumors and 14% of all primary brain and CNS tumors.^1^ The estimated annual incidence is 3.28 cases per 100,000 individuals.^1^ Diagnostic and prognostic stratification are guided by molecular markers such as isocitrate dehydrogenase mutation status and O-6-methylguanine DNA methyltransferase (MGMT) promoter methylation.^2,3^ Standard treatment for GBM includes maximal safe surgical resection followed by concurrent chemoradiotherapy with temozolomide (TMZ) and adjuvant TMZ therapy.^3^ Yet median overall survival (OS) remains approximately 21 months with the addition of tumor-treating fields.^4^ Immunotherapies and targeted agents have so far translated into limited and variable benefit. ^5–7^ Treatment is therefore life-prolonging rather than curative, and the biological reasons for this therapeutic ceiling remain poorly understood.

One aspect that has received comparatively little attention in this context is the microbiome. In other malignancies, host-microbiome interactions have moved from association to demonstrated mechanism: gut microbiome composition determines primary resistance to programmed cell death protein 1 blockade in lung and renal cancer, to the extent that antibiotic exposure reduces clinical benefit,^8^ and intratumoral bacteria can inactivate a chemotherapeutic drug enzymatically.^9^ Altered gut microbiota composition has been described across cancer types, most extensively in colorectal cancer, where it modulates inflammation and immune surveillance,^10^ although the mechanisms remain incompletely resolved.^11^ Unlike almost every tumor-intrinsic feature, the microbiota is in principle modifiable, which raises the question of whether comparable interactions contribute to the therapeutic resistance seen in GBM.

The gut-brain axis (GBA) provides a biologically plausible conduit linking intestinal microbes to CNS immunity.^12^ It operates through neural, hormonal, and immunological pathways carrying information in both directions.^13^ Its clinical relevance is best documented outside oncology: irritable bowel syndrome, inflammatory bowel disease (IBD), and neuropsychiatric conditions such as depression and anxiety have all been associated with disruptions of this axis.^14–16^ Across these conditions, inflammation and immune activation recur as a common mechanistic theme.

The GBA has more recently been proposed as a modulator of GBM development, progression, and treatment response, with microbial metabolites, systemic immune modulation and barrier permeability suggested as underrecognized contributors to immune dysregulation and tumor-promoting inflammation.^17–20^ Preclinical studies support this: mouse models show that the gut microbiota can influence the efficacy of TMZ through immune-mediated pathways.^21^ Upregulation of zonulin, which regulates epithelial and blood-brain barrier permeability, has been implicated in creating a permissive environment for tumor progression.^22^ Translational and clinical studies remain scarce. A recent phase I/II clinical trial reported that a high transcriptome-derived immune infiltration score, itself associated with higher fecal alpha diversity, predicted improved survival in GBM patients treated with anti-programmed-cell-death-ligand 1 therapy, underscoring the potential of microbiome-informed personalization of immunotherapy.^23^ It remains one of very few studies to examine GBA-related parameters directly in human GBM patients and the descriptive groundwork is still largely missing.

Bacteria dominate microbiome literature, but they are not the only microbial residents of the gut. Fungi account for only a small fraction of intestinal sequencing reads,^24^ yet conserved cell wall components such as β-glucans are recognized by the Dectin-1 receptor, triggering pro-inflammatory cytokine production.^25^ In mice, mucosa-associated fungi maintain epithelial barrier function and influence social behavior through type 17 immunity,^26^ and systemically administered β-glucan can train microglial responses.^27^ In humans, an expansion of intestinal CTG-clade fungi, which can elicit Th17 responses, has been associated with fibrosis progression in metabolic dysfunction-associated steatotic liver disease.^28^ In oncology, pan-cancer analyses have described tumor-type-specific fungal signatures across human malignancies.^29,30^ In GBM, the gut mycobiome has to our knowledge not been characterized, and no study has assessed both kingdoms in the same patients.

We conducted a prospective, monocentric case-control study to determine whether the gut microbiome of patients with newly diagnosed GBM differs from that of healthy controls, and whether such differences relate to circulating cytokine levels, the intratumoral myeloid compartment, or OS. An additional objective was to establish whether such a design is feasible at all in a severely affected patient cohort. We characterized both bacterial and fungal gut microbiota and identified differentially abundant taxa between patients and controls and, within the patient cohort, between patients with shorter and longer OS. To investigate potential immune-mediated mechanisms, we measured peripheral cytokine levels in both patients and controls and quantified myeloid cell markers in tumor tissue. We further examined whether the intratumoral myeloid compartment was associated with OS after adjustment for established clinical prognostic factors. Together, these analyses aim to provide insight into microbiome-related signatures and their potential relevance for GBM pathophysiology and clinical management.

## Results

### GBM patients and healthy controls are demographically comparable but differ in comorbidity and co-medication

Between October 2021 and August 2024, we prospectively recruited 23 patients with newly diagnosed, primary GBM and 23 healthy controls at the Department of Neurosurgery, University Hospital Würzburg. Patients were slightly older than controls (median 65 compared to 59 years) and predominantly male (60.9%), whereas the controls were slightly more often female (56.5%) (Table 1). The median Karnofsky performance status (KPS) at diagnosis was 90 and MGMT promoter methylation was present in 21.7% of patients (Table 2). All patients underwent surgery, most commonly subtotal resection (47.8%). An overview of participant recruitment, sample availability, and analyses is presented in Figure 1. The two groups were thus broadly comparable in demographic terms but differed, as expected for a tumor cohort, in comorbidity and co-medication, most notably corticosteroids and gastric protection.

**Figure 1:**
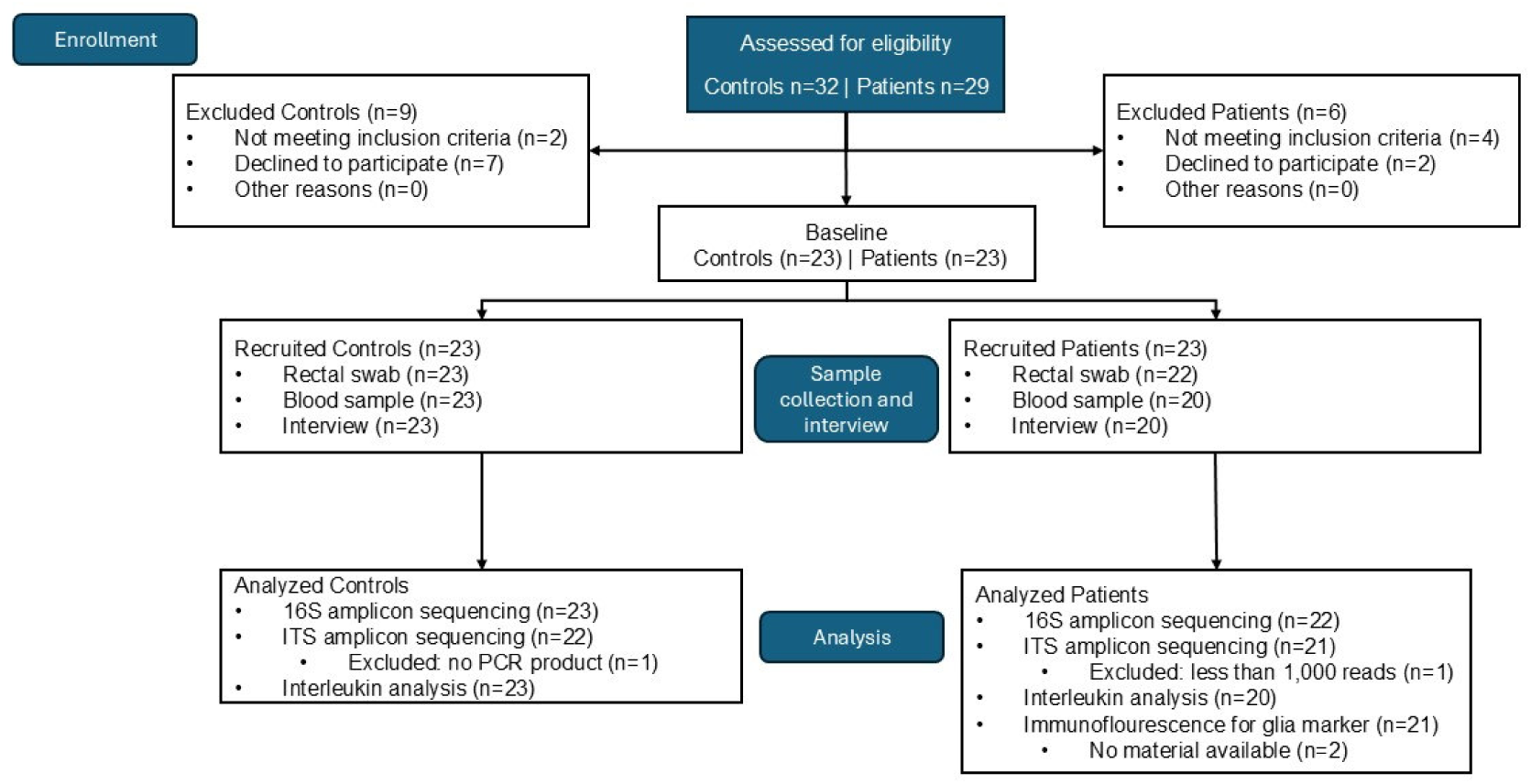
Study flow diagram. The diagram shows enrolment, exclusions, and the number of controls and GBM patients contributing to 16S amplicon sequencing, ITS amplicon sequencing, the interleukin panel and for the patients the immunofluorescence staining. Sample availability differed between modalities.

**Table 1:** Characteristics of study participants. Numerical variables are shown as median [interquartile range], categorical variables as number (percentage). Percentages refer to participants with available data. CREA, creatinine; CRP, C-reactive protein; GFR, glomerular filtration rate; GOT, glutamic oxaloacetic transaminase; GPT, glutamic pyruvic transaminase. ^a^ available for 20 patients; ^b^ available for 22 controls; ^c^ available for 21 patients.

| Variable | Controls (n=23) | Patients (n=23) | Total (n=46) |
| --- | --- | --- | --- |
| Age | 59.0 [55.5-66.5] | 65.0 [62.0-70.0] | 63.5 [57.0-69.0] |
| Sex: female | 13 (56.5%) | 9 (39.1%) | 22 (47.8%) |
| Cigarettes (/day) | 0.0 [0.0-1.5] | 0.0 [0.0-0.0] | 0.0 [0.0-0.0] |
| Alcohol (g/day)<sup>a</sup> | 12.5 [7.9-22.6] | 7.1 [2.7-18.5] | 10.9 [5.8-22.6] |
| Meat Consumption<sup>a</sup>: |  |  |  |
| none | - | 1 (5.0%) | 1 (2.3%) |
| several per month | 6 (26.1%) | 1 (5.0%) | 7 (16.3%) |
| several per week | 15 (65.2%) | 11 (55.0%) | 26 (60.5%) |
| daily | 2 (8.7%) | 7 (35.0%) | 9 (20.9%) |
| Carbohydrates Consumption<sup>a</sup>: |  |  |  |
| several per month | 2 (8.7%) | - | 2 (4.7%) |
| several per week | 5 (21.7%) | 2 (10.0%) | 7 (16.3%) |
| daily | 16 (69.6%) | 18 (90.0%) | 34 (79.1%) |
| Plants Consumption<sup>a</sup>: |  |  |  |
| none | 4 (17.4%) | 10 (50.0%) | 14 (32.6%) |
| irregular | 3 (13.0%) | 5 (25.0%) | 8 (18.6%) |
| several per month | 9 (39.1%) | 2 (10.0%) | 11 (25.6%) |
| several per week | 5 (21.7%) | 2 (10.0%) | 7 (16.3%) |
| daily | 2 (8.7%) | 1 (5.0%) | 3 (7.0%) |
| Probiotics Frequency<sup>a</sup>: |  |  |  |
| none | 5 (21.7%) | 1 (5.0%) | 6 (14.0%) |
| irregular | 4 (17.4%) | - | 4 (9.3%) |
| several per month | 9 (39.1%) | 5 (25.0%) | 14 (32.6%) |
| several per week | 4 (17.4%) | 8 (40.0%) | 12 (27.9%) |
| daily | 1 (4.3%) | 6 (30.0%) | 7 (16.3%) |
| Diabetes: |  |  |  |
| no | 23 (100.0%) | 19 (82.6%) | 42 (91.3%) |
| not insulin dependent | - | 4 (17.4%) | 4 (8.7%) |
| Blood Glucose (mg/dl) | 102.0 [91.0-130.5] | 105.0 [93.0-131.5] | 104.0 [91.2-132.8] |
| CREA (mg/dl) | 0.9 [0.8-1.0] | 0.9 [0.7-1.1] | 0.9 [0.7-1.0] |
| GFR<sup>b</sup> (ml/min) | 89.0 [77.8-99.8] | 79.0 [70.5-85.0] | 82.0 [75.0-92.0] |
| GOT (U/l) | 22.6 [20.8-28.0] | 24.8 [18.5-34.0] | 23.1 [19.9-28.0] |
| GPT<sup>c</sup> (U/l) | 21.9 [17.1-26.0] | 29.1 [21.3-56.3] | 23.4 [19.9-36.7] |
| CRP (mg/dl) | 0.0 [0.0-0.1] | 0.0 [0.0-0.3] | 0.0 [0.0-0.2] |
| Antihypertensives | 6 (26.1%) | 15 (65.2%) | 21 (45.7%) |
| Anticoagulants | 1 (4.3%) | 11 (47.8%) | 12 (26.1%) |
| Statins | 1 (4.3%) | 6 (26.1%) | 7 (15.2%) |
| Antidiabetics | - | 4 (17.4%) | 4 (8.7%) |
| Gastric Protection | 1 (4.3%) | 16 (69.6%) | 17 (37.0%) |
| Analgesics | - | 5 (21.7%) | 5 (10.9%) |
| Antidepressants | 2 (8.7%) | 1 (4.3%) | 3 (6.5%) |
| Sedatives | - | 2 (8.7%) | 2 (4.3%) |
| Thyroid | 1 (4.3%) | 2 (8.7%) | 3 (6.5%) |
| Hormonal | - | 2 (8.7%) | 2 (4.3%) |
| Prostate | - | 3 (13.0%) | 3 (6.5%) |
| Corticosteroids | - | 11 (47.8%) | 11 (23.9%) |
| Antiepileptics | - | 4 (17.4%) | 4 (8.7%) |

**Table 2:**
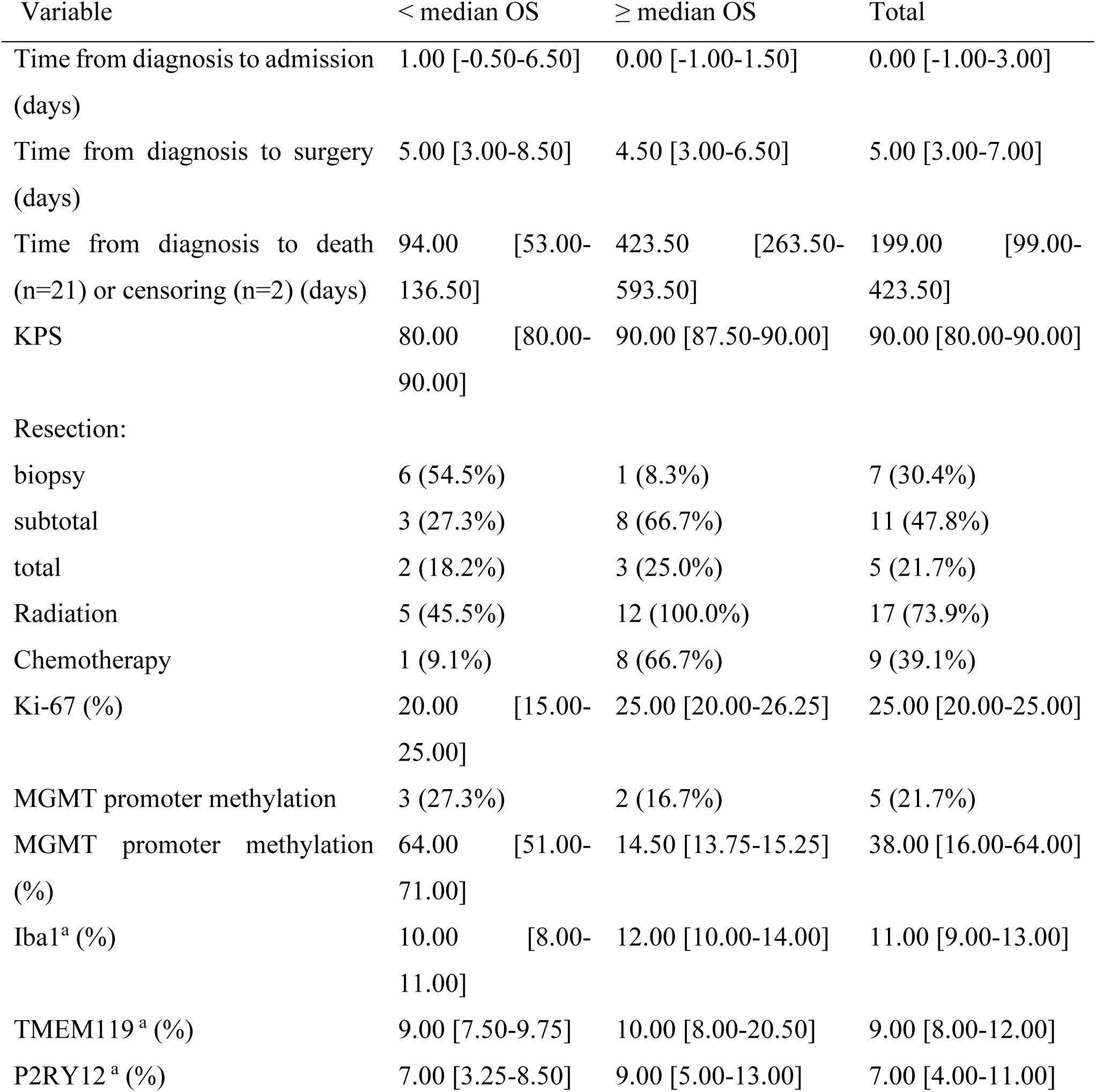
Clinical and histopathological characteristics of GBM patients. Patients were grouped by overall survival (OS), split at the cohort median OS (< median OS, n = 11; ≥ median OS, n = 12). Numerical variables are shown as median [interquartile range], categorical variables as number (percentage). Negative values for time from diagnosis to admission indicate admission before diagnosis. MGMT promoter methylation (%) is given for tumors with MGMT promoter methylation. Iba1, TMEM119 and P2RY12 are given as percentage of DAPI-positive nuclei. DAPI, 4′,6-diamidino-2-phenylindole; GBM, glioblastoma; Iba1, ionized calcium-binding adapter molecule 1; Ki-67, proliferation marker Ki-67; KPS, Karnofsky performance status; MGMT, O-6-methylguanine-DNA methyltransferase; P2RY12, purinergic receptor P2Y12; TMEM119, transmembrane protein 119. ^a^ available for 21 patients.

### Reduced fungal alpha diversity in GBM patients

We characterized the gut bacteriome and mycobiome from rectal swabs, sequencing the 16S rRNA gene and the internal transcribed spacer (ITS) region so that both kingdoms could be assessed. After quality filtering, 16S sequencing yielded 2,994,178 reads across 45 samples (23 controls, 22 patients, median 65,605 reads per sample) and ITS sequencing 963,388 reads across 43 samples (22 controls, 21 patients, median 12,147), after exclusion of two samples with no PCR product or fewer than 1,000 reads. Agglomeration yielded 218 bacterial and 102 fungal taxa.

The two communities were very different in evenness. The bacterial community was dominated by *Prevotella* (mean relative abundance 8.7%), *Bacteroides* (6.7%), *Phocaeicola* (6.6%), *Finegoldia* (5.8%) and *Pseudescherichia* (4.9%), followed by *Ruminococcus*, *Faecalibacterium*, *Corynebacterium* and *Blautia* (3.2-3.8% each) (Figure 2A). The fungal community was far less even: *Debaryomyces hansenii* (37.9%) and *Saccharomyces cerevisiae* (19.8%) together accounted for more than half of all fungal reads, followed by *Nakaseomyces glabratus* (4.5%; formerly *Candida glabrata*), unclassified Malasseziaceae (4.4%) and a set of taxa with relative abundance below 3% each (Figure 2B). Two features warrant caution. *D. hansenii* was also the dominant taxon in the ITS buffer control, so its apparent abundance cannot be cleanly separated from a reagent-derived signal. Several other abundant taxa (*Fomitopsis pinicola*, *Trametes versicolor*, *Phlebia bresadolae*, unclassified Orbiliaceae) are wood-decay or soil-associated fungi without a described intestinal niche, and most plausibly represent dietary or environmental input.

**Figure 2.**
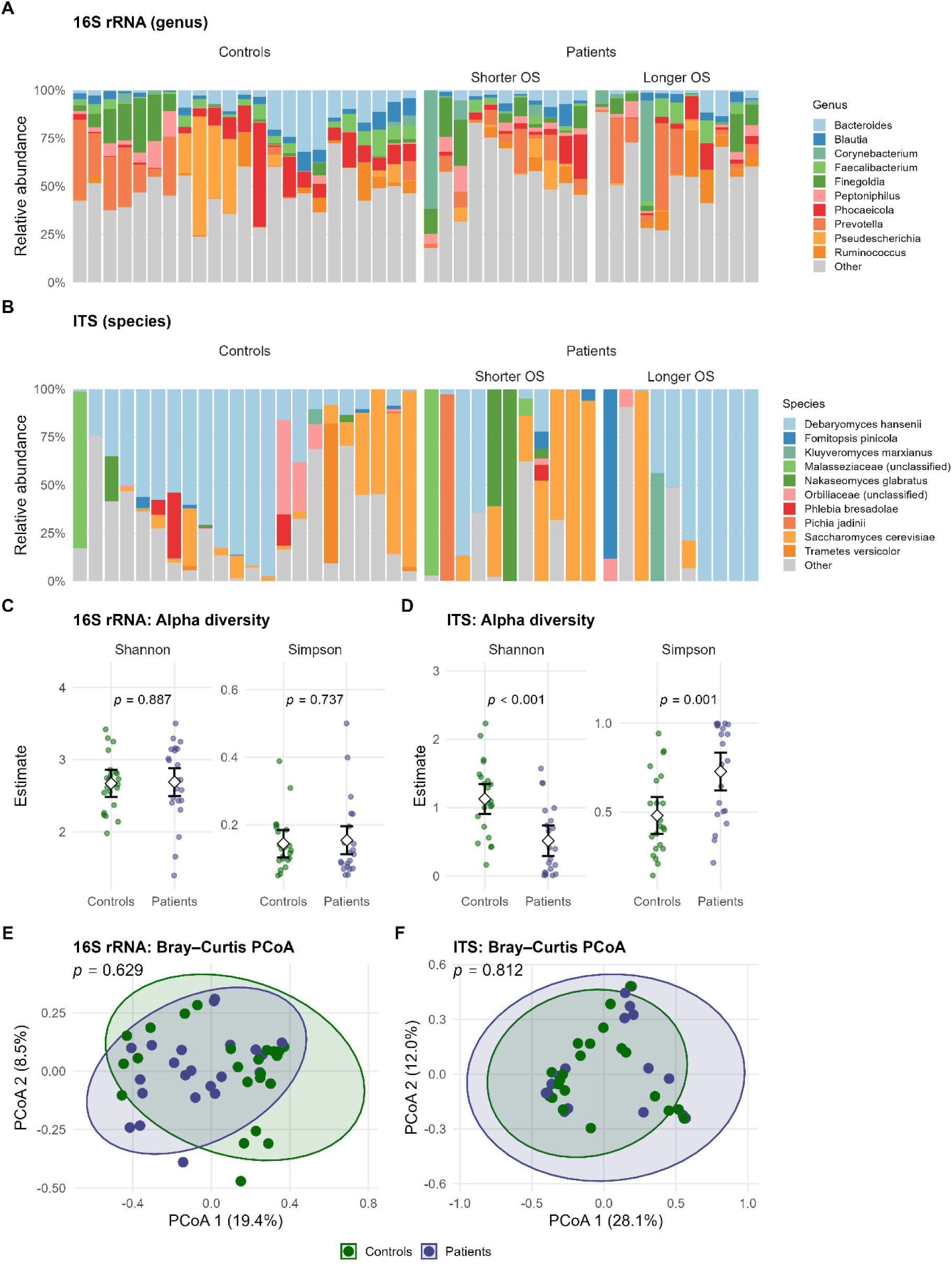
Gut bacterial and fungal communities in GBM patients and healthy controls. **(A, B)** Relative abundance of the ten most abundant bacterial genera (A, 16S rRNA) and fungal species (B, ITS); remaining taxa are grouped as “Other”. Each bar is one participant, ordered by Bray-Curtis clustering within groups. **(C, D)** Shannon and Simpson diversity for 16S (C) and ITS (D). Small points show per-sample DivNet estimates, diamonds and error bars the group means with 95% confidence intervals estimated by betta; p-values from betta. **(E, F)** Principal coordinates analysis of Bray-Curtis dissimilarities for 16S (E) and ITS (F); ellipses indicate 95% confidence regions, axis labels the variance explained.

To characterize the communities as a whole, we compared alpha and beta diversity between patients and controls (Figure 2C-F). Fungal communities were less diverse in patients, with a lower Shannon index (estimated mean 0.51 versus 1.13; p < 0.001) and a higher Simpson index (Σpᵢ², where higher values indicate stronger dominance by few taxa; 0.73 versus 0.48; p = 0.001) (Figure 2D). The bacteriome showed no such shift (Shannon p = 0.887; Simpson p = 0.737; Figure 2C). Overall community composition, assessed by Bray-Curtis dissimilarity, did not differ for either kingdom (16S p = 0.629; ITS p = 0.812), and the principal coordinate analyses showed largely overlapping distributions (Figure 2E, F).

### Depletion of Lachnospiraceae and Ruminococcaceae and increase of *Methanobrevibacter* and *Nakaseomyces glabratus* in GBM patients

To examine whether individual taxa differed between the groups, all genera and species were tested for differential abundance between patients and controls using robust score tests, without covariates given the limited sample size. This exploratory analysis identified 14 of 218 bacterial and archaeal taxa and 4 of 102 fungal species as differing at a nominal, uncorrected threshold of p ≤ 0.05. Twelve of the bacterial and archaeal taxa were resolved to genus level; two could not be resolved beyond family and are reported but not interpreted further.

The bacterial/archaeal signal was markedly asymmetric and concentrated in a few clades: eleven of the fourteen taxa were more abundant in controls (Figure 3A). Four belonged to the Lachnospiraceae: *Anaerostipes* (log-fold change −1.61, p = 0.017), *Fusicatenibacter* (−1.50, p = 0.025), *Coprococcus* (−0.90, p = 0.050) and an unclassified Lachnospiraceae group (−1.16, p = 0.022); and two each to the Ruminococcaceae (*Agathobaculum*, −1.44, p = 0.001; *Flintibacter*, −1.00, p = 0.020), the Erysipelotrichaceae (*Faecalibacillus*, −2.86, p = 0.038; *Holdemania*, −1.00, p = 0.021) and the Eggerthellaceae (*Adlercreutzia*, −1.20, p = 0.015; unclassified, −0.54, p = 0.048), alongside *Bacteroides* (−1.09, p = 0.047). Only three taxa showed the opposite direction, being more abundant in patients: the methanogenic archaeon *Methanobrevibacter* (+2.10, p = 0.024), *Schaalia* (+1.90, p = 0.017) and *Negativibacillus* (+1.83, p = 0.015). All fourteen were detected in at least 20 of the 45 samples.

**Figure 3.**
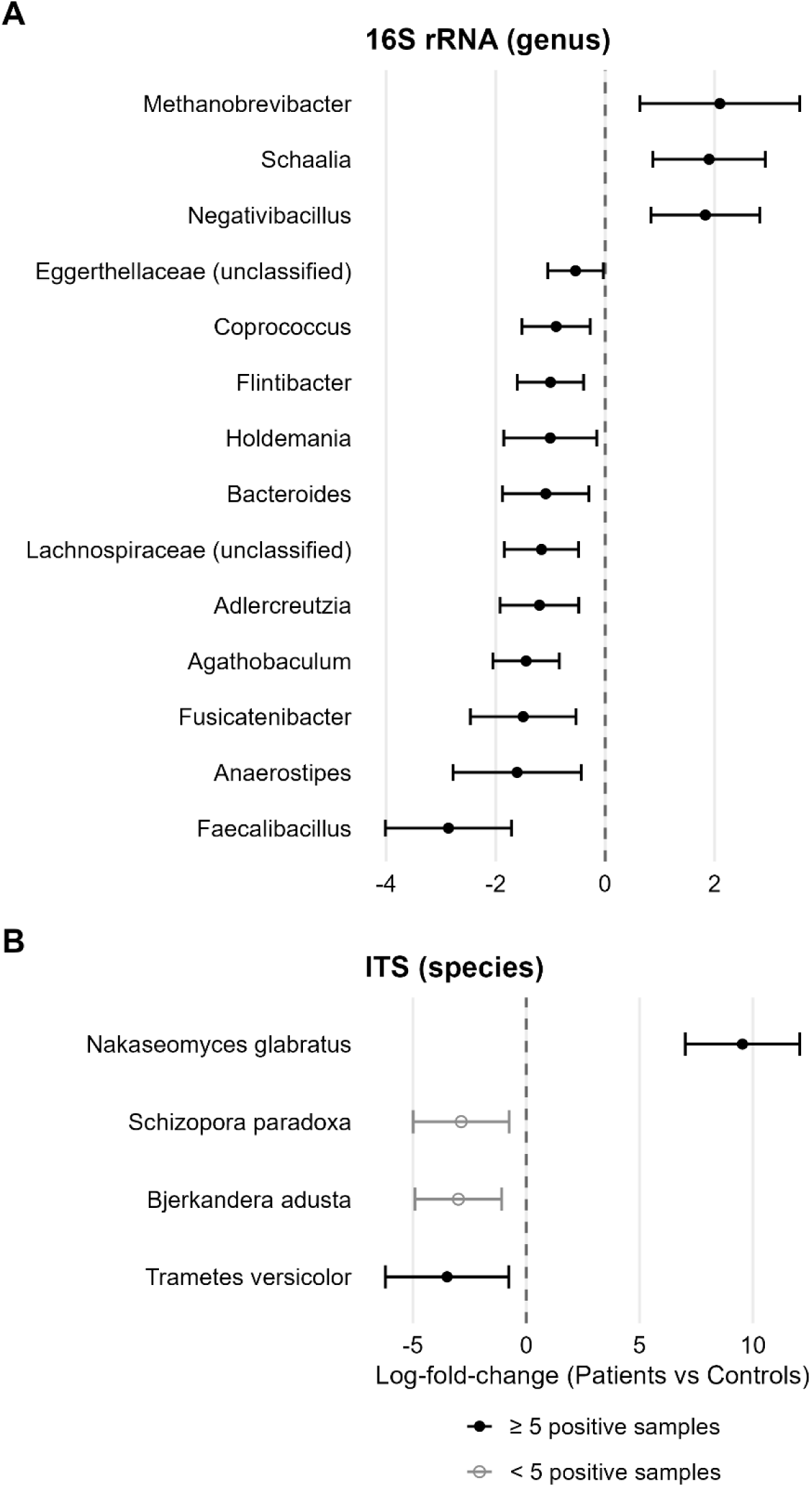
Taxa differing in abundance between GBM patients and healthy controls. Log-fold-changes with 95% confidence intervals for **(A)** bacterial genera and **(B)** fungal species; positive values indicate higher abundance in patients. All taxa with an uncorrected p ≤ 0.05 are shown; open symbols mark taxa detected in fewer than five samples, for which estimates are less stable. The dashed line marks no difference.

The fungal contrast was smaller (Figure 3B). *N. glabratus* (formerly *C. glabrata*) showed a higher estimated abundance in patients (+9.54, p = 0.041) but was detected in only 13 of 43 samples and has a wide 95% confidence interval (7.02-12.07). The three taxa with higher estimates in controls, *T. versicolor* (−3.49, p = 0.003), *B. adusta* (−2.99, p = 0.047) and *S. paradoxa* (−2.87, p = 0.025), are wood-decay fungi detected in 6, 2 and 4 samples respectively, and thus correspond to the environmentally derived fraction described above. Neither of the two dominant fungal taxa differed significantly between groups (*D. hansenii*, +5.51, p = 0.062; *S. cerevisiae*, +5.74, p = 0.066).

None of these taxa retained significance after Benjamini-Hochberg correction, for either marker. Estimates, confidence intervals and adjusted p-values for all tested taxa are provided in Supplementary Data 1.

Taken together, these analyses identify a small set of candidate taxa that distinguish GBM patients from controls. They are concentrated in a few clades and consistent in direction.

### Limited evidence for a systemic inflammatory signature

To assess whether the gut microbial differences described above were accompanied by a systemic inflammatory signature, we quantified plasma concentrations of seven cytokines (IFN-α, IL-2, IL-4, IL-6, IL-10, IL-17A/F and TNF-α) by multiplex immunoassay in 20 patients and 23 controls. Concentrations were compared between groups by Wilcoxon rank-sum tests. IL-10 showed the smallest p-value without reaching statistical significance (p = 0.06) and was quantifiable in 3 of 20 patients but in none of the 23 controls (Figure 4A). IL-17A/F and IL-2 showed the same tendency (IL-17A/F, p = 0.14; IL-2, p = 0.13) and were likewise more frequently quantifiable in patients than in controls (4 of 20 versus 1 of 23; 8 of 20 versus 5 of 23). The remaining analytes showed no such tendency. Given that most concentrations were at or below the limit of detection, IL-10, IL-17A/F and IL-2 are reported as candidates for more sensitive assays rather than as established differences.

**Figure 4.**
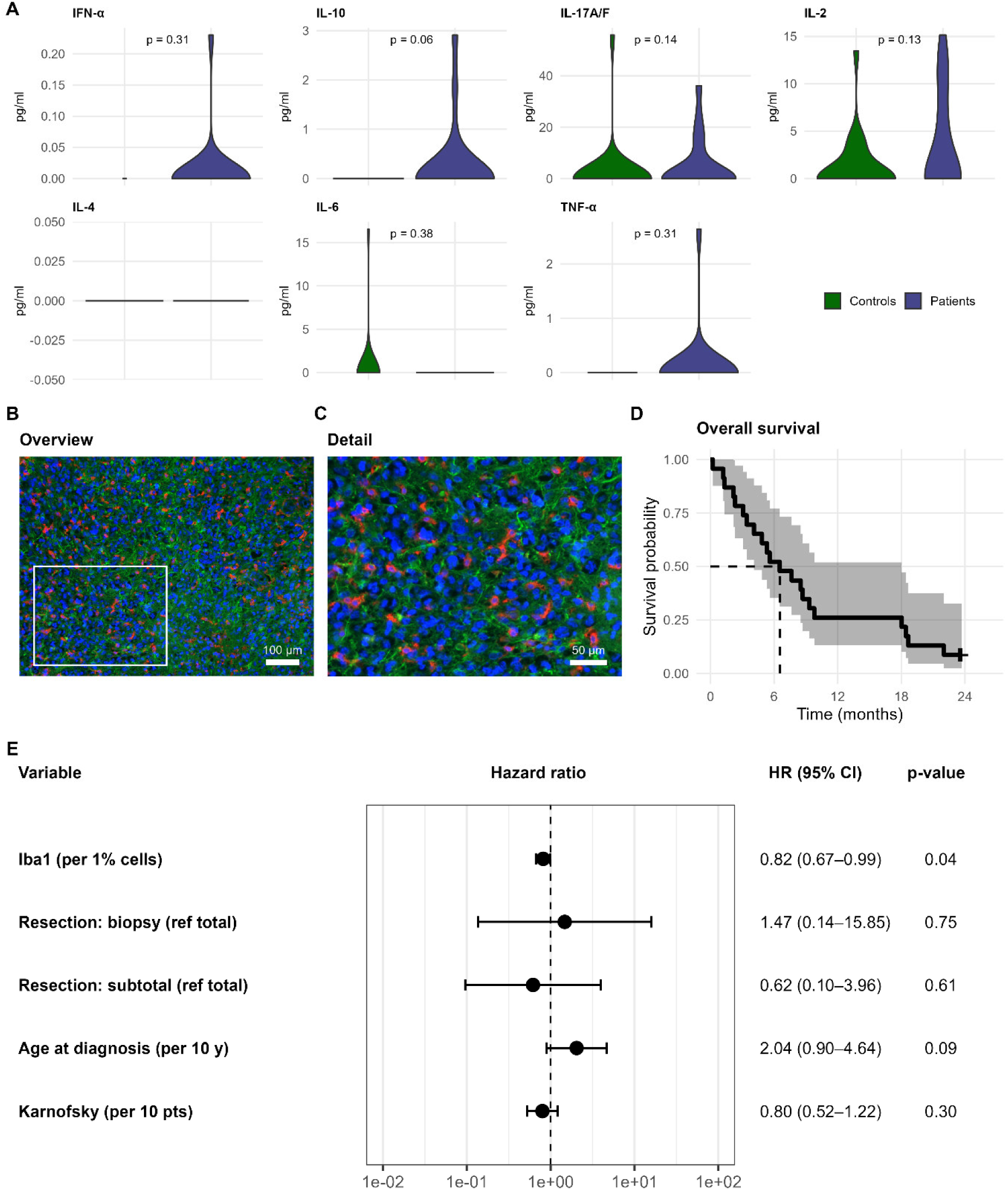
Systemic and intratumoral immune profile and association with OS. **(A)** Plasma concentrations of seven cytokines in patients and controls; p-values from Wilcoxon rank-sum tests. **(B, C)** Representative immunofluorescence staining demonstrating microglial distribution within GBM (Iba1 red, GFAP green, DAPI blue). The rectangle in (B) marks the region enlarged in (C). Scale bars, 100 µm (B) and 50 µm (C). **(D)** Kaplan-Meier estimate of OS for the whole cohort (n = 23); shaded area 95% confidence interval, tick marks censored observations. **(E)** Multivariable Cox model. Hazard ratios with 95% confidence intervals refer to a 1% increase in Iba1 immunoreactivity, a 10-year increase in age and a 10-point increase in KPS; extent of resection (total, subtotal, biopsy).

### Iba1 density associated with longer overall survival

To characterize the local compartment, we stained tumor tissue, available for 21 of the 23 patients, for the myeloid markers ionized calcium-binding adapter molecule 1 (Iba1), transmembrane protein 119 (TMEM119) and purinergic receptor P2Y12 (P2RY12), using glial fibrillary acidic protein (GFAP) as a tumor-tissue reference for region selection. Iba1-positive cells were distributed throughout the tumor parenchyma, interspersed between GFAP-positive tumor cells rather than confined to focal aggregates (Figure 4B, C). Iba1-positive cells accounted for a median of 11% of all 4′,6-diamidino-2-phenylindole (DAPI)-positive nuclei (interquartile range; IQR 9-13%), the microglia-restricted markers TMEM119 and P2RY12 for 9% (IQR 8-12%) and 7% (IQR 4-11%), respectively (Table 2).

Median OS of all GBM patients was 6.5 months (199 days; 95% CI 4.1-18.0 months; Figure 4D); two patients were alive at the end of follow-up and were censored. The candidate tissue markers Iba1, TMEM119 and P2RY12 together with Ki-67 and MGMT promoter methylation were initially screened in univariate Cox models using the prespecified likelihood-ratio and AIC criteria. Only Iba1 met both selection criteria and was subsequently entered into the multivariable model together with age, KPS and extent of resection. None of the microglia-restricted markers, Ki-67 or MGMT promoter methylation did so; the latter was available in only five patients (Supplementary Table 1). In the final model, higher Iba1 immunoreactivity was associated with longer OS (HR 0.82 per percentage point, 95% CI 0.67-0.99, p = 0.04; Figure 4E). No confounder reached significance (age, HR 2.04 per 10 years, p = 0.09; KPS, HR 0.80 per 10 points, p = 0.30; extent of resection p > 0.6). The estimate was robust across sensitivity models with alternative adjustment sets (Supplementary Table 2). Given 19 events and five model parameters, however, this model is estimated on limited information, and the association should be regarded as exploratory.

### Few microbial differences between survival groups

We finally asked whether the microbial and immunological measurements were related to outcome within the patient cohort. We stratified patients at the cohort’s median OS into a shorter-survival (n = 11) and a longer-survival group (n = 12); the two patients alive at censoring had the longest follow-up and were assigned unambiguously to the longer-survival group. Neither bacterial nor fungal alpha diversity differed between the survival groups (16S: Shannon p = 0.493, Simpson p = 0.432; ITS: Shannon p = 0.576, Simpson p = 0.677; Supplementary Figure 1A, B).

Likewise, overall community structure was similar in both survival groups (Bray-Curtis, 16S, p = 0.67; ITS p = 0.51; Additional file 1, Supplementary Figure 1C, D). For 16S, 2 genera reached an uncorrected p ≤ 0.05, both at higher abundance in the shorter-survival group (Lawsonibacter, log-fold change −0.77, p = 0.03; Turicibacter, −1.12, p = 0.05). For ITS, 2 species reached this threshold: *D. hansenii*, at higher abundance in the longer-survival group (+4.04, p = 0.01), and an unclassified *Orbilia* taxon detected in only three samples (−2.36, p = 0.05). *D. hansenii* was, however, also detected at high levels in a negative control, so its abundance may reflect a reagent-derived signal. No taxon remained significant after Benjamini-Hochberg correction for either marker (Supplementary Figure 2, Supplementary Data 1).

Plasma cytokine concentrations did not differ between the survival groups for any of the seven analytes (Supplementary Figure 3).

## Discussion

In this prospective case-control study we characterized the gut bacteriome and mycobiome of patients with newly diagnosed GBM together with healthy controls. We also examined systemic cytokines, the intratumoral myeloid compartment and OS. At community level, only the mycobiome showed differences. Patients harbored a less diverse fungal community than controls. In contrast, bacterial diversity, overall community structure and diversity by survival group were similar. At the taxon level, several butyrate– and propionate-producing bacteria were nominally reduced in patients, while *Methanobrevibacter* and *N. glabratus* (formerly *C. glabrata*) were enriched; none of these differences withstood correction for multiple testing. In a separate analysis of the tumor tissue, a higher density of Iba1-positive myeloid cells within the tumor was associated with longer OS.

Bacterial diversity did not differ between patients and controls, suggesting that GBM at diagnosis is not associated with a strong shift in the overall bacterial community. Diversity also did not differ between survival groups. A prospective GBM cohort with survival data likewise found no significant difference.^23^ The lower fungal diversity in patients needs to be interpreted with care. Since the mycobiome was dominated by *D. hansenii* and *S. cerevisiae*, diversity indices respond strongly to changes in these two taxa and *D. hansenii* was also the dominant taxon in a buffer control. Additionally, acid suppression, which was more common in patients, promotes yeast colonization^31^ and may have contributed to a community dominated by fewer taxa.

Eleven of the fourteen nominally differential bacterial and archaeal taxa were less abundant in patients, belonging mainly to the Lachnospiraceae and Ruminococcaceae. The depleted genera include several recognized butyrate producers (*Agathobaculum*, *Anaerostipes*, *Flintibacter*, *Coprococcus*)^32–35^ and propionate producers (*Bacteroides*, *Adlercreutzia*).^32,36^ Short-chain fatty acids are the principal energy source of colonocytes, maintain epithelial barrier function,^37^ and act beyond the gut: in germ-free mice, microglia cells are immature and functionally impaired, and supplementation with short-chain fatty acids restores their number, morphology and function.^38^ A potentially reduced capacity for short-chain fatty acid production would therefore be expected to affect both the intestinal barrier and the myeloid compartment of the CNS. Such a shift is not specific to GBM. Taxa of this kind are consistently reported as reduced across chronic inflammatory, metabolic and neurological conditions, and a cross-disease meta-analysis found that they are associated with health in general rather than with any one disease.^39^ Among the enriched taxa, *Schaalia* is part of the oral microbiota,^40^ and its recovery from the lower gastrointestinal tract could reflect gastric protection, which was more prevalent in the GBM group and leads to an oralization of the fecal community.^41^ *Methanobrevibacter* was the only enriched taxon for which a cancer-associated role has been proposed. *M. smithii* is enriched in colorectal cancer across independent multi-cohort analyses.^42,43^ *In vitro* work indicates that it engages in mutualistic metabolic exchange with cancer-associated bacteria, including *Fusobacterium nucleatum*.^42^ *Methanobrevibacter* might therefore be an interesting candidate for further studies.

Of the four fungal species, *N. glabratus* (formerly *C. glabrata*) is the only one colonizing the gut and the only one enriched in patients. Acid suppression might be a possible confounder, as proton pump inhibitor use increases gastric yeast colonization.^31^ Antibiotic exposure, which is another established driver, was an exclusion criterion at recruitment. *N. glabratus* is therefore the fungal taxon in our data most likely to reflect a real change in the intestinal community. The three depleted species are wood-decaying fungi that cannot grow under gut conditions, and controlled-diet experiments indicate that much of the fungal DNA recovered from stool is transient and derived from the environment.^44^ Their depletion more plausibly marks altered diet or exposure than a property of the gut mycobiome.

The comparison by survival group yielded few differentially abundant taxa. Four reached an uncorrected p ≤ 0.05 and none remained significant after correction. The two fungal taxa are not interpretable: *D. hansenii* was also detected at high levels in a negative control, and the unclassified *Orbilia* taxon was present in only three samples. *Lawsonibacter* and *Turicibacter* were both more abundant in the shorter-survival group. For *Turicibacter*, the direction is consistent with a murine colitis model in which the genus was linked to a pro-inflammatory cascade acting through a guanidino metabolite and myeloid-differentiation-primary-response signaling, with downstream effects on microglial polarization.^15^ Whether this bears on our observation cannot be decided here, and with eleven and twelve patients per group these results have to be interpreted with caution.

Plasma cytokine concentrations did not differ significantly between patients and controls. Values were at or below the limit of detection for most participants, and IL-10, IL-17A/F and IL-2 were the only analytes tending towards higher values in patients. Almost half of the patients but none of the controls received corticosteroids at sampling, which suppress cytokine production^45^ and may have attenuated levels in the patient group. Concentrations also did not differ by survival group.

Iba1, also known as allograft inflammatory factor 1, is a marker of microglia and macrophages and reflects the myeloid compartment of the CNS.^46,47^ In GBM, tumor-associated microglia and macrophages are among the dominant non-neoplastic cell populations of the tumor microenvironment and modulate tumor progression, immune suppression, invasion, edema formation, and treatment resistance.^48^ In our cohort, higher Iba1 immunoreactivity was associated with improved OS, suggesting that the myeloid contexture may provide prognostic information beyond established clinical parameters. Iba1 staining detects both resident microglia and infiltrating macrophages and does not distinguish activation states. This matters because tumor-associated microglia and macrophages, especially M2-like phenotypes, have been linked to aggressive glioma biology and poor outcome,^49^ whereas other data suggest context-dependent immune functions of specific microglial subpopulations.^50^ A larger Iba1-positive compartment may therefore reflect a more pronounced local immune surveillance, a less exhausted tumor microenvironment, or a phenotype more permissive to myeloid cell infiltration. Given the cohort size, the association is hypothesis-generating and requires validation with spatially resolved and phenotypically refined immune profiling.

Experimental work indicates that gut microbial composition influences microglial maturation and activation,^38^ which places the myeloid compartment at the interface between the microbial and tissue-level findings of this study. Establishing whether such a relationship exists in GBM will require larger cohorts combined with spatial immune phenotyping and metabolomics.

A further objective of this study was to assess whether prospective microbiome research is feasible in this patient population. It is, but requires adaptation to the circumstances of newly diagnosed GBM patients, who frequently face neurological deficits, cognitive impairment, and the burden of the diagnosis itself, so that procedures unproblematic in healthy individuals become demanding in routine clinical practice. Rectal swabs were initially intended to be self-collected on the evening before surgery, as in controls, but despite written and graphical instructions many patients reported feeling overwhelmed by this additional task. The protocol was therefore amended, and swabs were subsequently taken by trained personnel immediately after induction of anesthesia and before any surgical intervention. This improved acceptance, enabled reliable sample acquisition, and a biologically relevant alteration of the gut microbiome within minutes of induction is unlikely. It nonetheless introduced the difference in sampling conditions discussed above.

The modest cohort size is a further limitation, although it follows from the strict eligibility criteria. Participants with recent infections or vaccinations, antibiotic exposure, probiotic supplementation, autoimmune disorders, IBD, severe organ dysfunction, or previous malignancies were excluded, and recruitment took place during the COVID-19 pandemic, which further reduced the eligible pool. These criteria limited recruitment but removed major confounders. The monocentric design limits generalizability and warrants validation in independent cohorts. The median OS of 6.5 months was also shorter than the approximately 21 months reported for standard treatment,^4^ which likely reflects the composition of the cohort: 30.4% of patients underwent biopsy only and 47.8% subtotal resection, and MGMT promoter methylation, a predictor of benefit from temozolomide, was present in 21.7% of tumors compared with approximately 40% in other cohorts.^51^ The survival analyses therefore refer to a population with comparatively poor prognosis and may not transfer to patients with more favorable clinical profiles. Within these constraints, prospective recruitment, standardized clinical phenotyping, microbiome sequencing, cytokine profiling, and tissue-based analyses could be combined within the routine workflow of a tertiary neuro-oncology center.

This study contributes some of the first paired bacteriome and mycobiome data from newly diagnosed GBM patients and healthy controls. Among the taxa that differed between groups, the depletion of butyrate and propionate producers is the finding we consider most likely to carry forward. Short-chain fatty acids are established regulators of microglial maturation and function. Of the myeloid markers assessed, Iba1 density was independently associated with survival. We therefore hypothesize that the myeloid compartment is the interface at which the gut-level and tissue-level findings converge.

## Methods

### Study design

Patients with radiomorphological suspicion of GBM scheduled for surgical resection or biopsy were informed about the study and screened against predefined criteria after written informed consent. In consenting patients, a rectal swab was collected at the time of anesthesia induction and an additional 10 ml of Ethylenediaminetetraacetic acid (EDTA) blood was drawn alongside routine preoperative blood sampling. Patients were formally included in the study upon histopathological confirmation of GBM, and clinical and histopathological data were documented on an internal report form. Intraoperatively resected tumor tissue was formalin-fixed, paraffin-embedded for immunofluorescence staining, without interfering with routine neuropathological diagnostics. A structured interview was conducted postoperatively to assess lifestyle and dietary habits.

Control participants were enrolled at the outpatient neurosurgery clinic. After verbal and written informed consent, 10-25 ml of EDTA blood was drawn, a rectal swab was self-collected, and the structured interview was conducted on the day of study inclusion. All plasma tubes and rectal swabs from study participants were cryopreserved at –80 °C.

### Patient and participant inclusion

All patients were treated at the Department of Neurosurgery, University Hospital Würzburg, Germany, between October 2021 and August 2024. Healthy control participants from the same catchment area were recruited through public postings on institutional bulletin boards and via a study information flyer. Controls were not individually matched to patients; age and sex distributions were compared between groups. Inclusion followed the declaration of Helsinki, and as approved by the Institutional Review Board of the University of Würzburg (# 16-/21-am). Eligible patients were aged over 18 years, showed radiomorphological suspicion of GBM and were competent to provide informed consent. Histopathology was validated by a neuropathologist (CMM) according to the 2021 WHO criteria.^2^ Exclusion criteria were histologically confirmed gliomas of CNS WHO grade 1, 2 or 3; a non-malignant intracranial mass or cerebral metastasis of a different tumor; heart failure of New York Heart Association class III/IV; renal insufficiency of grade 4 or 5; liver insufficiency of Child-Pugh class B/C; obesity with a body mass index above 35 kg/m2; chronic inflammatory bowel disease; rheumatic or autoimmune disease; any malignant disease within the preceding ten years; infection within four weeks before enrollment; antibiotic use within the preceding six months; vaccination within four weeks before enrollment; intake of probiotics in enteric-coated capsules; and pregnancy. Healthy controls were adults (≥18 years) capable of providing informed consent, with the same exclusion criteria plus any clinical or histological evidence of structural CNS disease and recent corticosteroid use within four weeks before enrollment. No formal sample size calculation was performed. The sample size was determined by the number of eligible participants recruited during the study period.

### Questionnaire

A structured questionnaire assessed lifestyle and dietary habits over the preceding six months. Data was collected digitally using EvaSys (version V9.1 2459) and Microsoft Excel (version 2409). The questionnaire was completed in a structured interview format and assessed smoking, alcohol consumption, and dietary habits, including the intake of meat, carbohydrates, plant-based meals, and probiotics. Particular focus was placed on factors known to affect the gut microbiome, including antibiotic use (≤6 months), recent vaccination (≤4 weeks), recent infection (≤4 weeks), and gastrointestinal disease. For patients, the interview was typically conducted two to three days postoperatively, in the control group, interviews were performed on the day of study inclusion. Consumption in each dietary category was recorded on a five-level ordinal frequency scale (none, irregular, several times per month, several times per week, daily). Average daily alcohol intake (g/day) was estimated by combining the reported drinking frequency and quantity with beverage-specific mean alcohol contents per litre,^52^ and the number of cigarettes per day was taken directly from the questionnaire, with non-smokers coded as zero.

### DNA isolation

Rectal swabs (Copan FLOQSwabs, Mast Group, Reinfeld, Germany) were stored at –80 °C. Swab material was transferred to microcentrifuge tubes using a custom-made filtration setup. Microbial DNA was isolated with the DNeasy PowerSoil Pro Kit (QIAGEN, Hilden, Germany), following the manufacturer’s instructions, except that each lysate was split across two spin columns, and pooled at elution to increase DNA yield. DNA was quantified on a Qubit 4 Fluorometer (dsDNA HS Assay Kit, Invitrogen, Thermo Fisher Scientific, Darmstadt, Germany) and stored at –20 °C until sequencing.

### Cytokine Analysis

Peripheral blood in EDTA tubes was left standing at room temperature for one hour, centrifuged at 1,500 × g for 10 minutes and the resulting plasma stored at –80 °C. Cytokines were measured with the ProcartaPlex™ Panel (Invitrogen, Thermo Fisher Scientific, Darmstadt, Germany), containing interferon-α (IFN-α), interleukin-2 (IL-2), IL-4, IL-6, IL-10, IL-17A/F, and tumor necrosis factor-α (TNF-α), according to the manufactureŕs instructions.

### Immunofluorescence

Formalin-fixed, paraffin-embedded tumor tissue was available for all but two patients (n = 21). For each tissue block, three separate 2.5 µm sections were stained for glial fibrillary acidic protein (GFAP), which served as a tumor-tissue reference for region selection and was not quantified, in combination with either ionized calcium-binding adapter molecule 1 (Iba1), transmembrane protein 119 (TMEM119) or purinergic receptor P2Y12 (P2RY12). Sections were deparaffinized, followed by heat-induced antigen retrieval in 10 mM citrate buffer for 20 min. After two washes of 5 min each in Tris-buffered saline with 0.1% Tween 20 (TBS-T), sections were blocked with 10% goat serum (Invitrogen, Thermo Fisher Scientific, Darmstadt, Germany). Primary antibodies were diluted in antibody dilution buffer (DCS Innovative Diagnostik-Systeme, Hamburg, Germany) and incubated overnight at 4 °C: GFAP (Santa Cruz Biotechnology, Dallas, TX, USA; sc-33673; 1:100), Iba1 (FUJIFILM Wako Pure Chemical Corporation, Osaka, Japan; 019-19741; 1:1000), TMEM119 (Abcam, Cambridge, UK; ab185333; 1:800) and P2RY12 (antibodies-online, Aachen, Germany; ABIN1387659; 1:200). Sections were washed twice for 5 min in TBS-T and incubated for 1 h at room temperature with Alexa Fluor Plus 555 goat anti-rabbit IgG (Invitrogen, Thermo Fisher Scientific, Darmstadt, Germany; A32732; 1:1000) and Alexa Fluor Plus 488 goat anti-mouse IgG (Invitrogen; A32723; 1:1000). After two further washes in TBS-T of 5 min each, sections were mounted in medium containing 4′,6-diamidino-2-phenylindole (DAPI; Abcam, Cambridge, UK).

Immunofluorescence images were acquired using the Leica DMI3000 B microscope at 10× magnification. Whenever possible, four non-overlapping regions of each tissue sample were imaged and analyzed. DAPI-positive nuclei were quantified automatically in Fiji (version 2.16.0/1.54p)^53^ using the StarDist plugin (version 0.3.0)^54^ with the pretrained 2D model Versatile (fluorescent nuclei). Default settings were used except for a probability threshold of 0.48 and an overlap threshold of 0.3. Marker-positive cells were counted manually on the same image fields, and the proportion of marker-positive cells per field was calculated against the StarDist-detected nuclei. Per-field counts were pooled across all evaluable fields of a section and are reported as the percentage of positive cells. Between two and four fields were evaluable per tumor.

### Metadata processing and descriptive analysis

Clinical metadata and medication data were imported from separate files and merged on the participant identifier; variables recorded in more than one file were checked for agreement and duplicates removed. Plasma cytokine measurements and myeloid marker quantifications (Iba1, TMEM119, P2RY12) were joined to the same table. All date variables were converted into intervals relative to the date of histopathological diagnosis (days from diagnosis to admission, to sample collection, and to death or last follow-up), and age at diagnosis was derived from the date of birth. ^69,70^ OS was defined as the time from diagnosis to death from any cause. Patients alive at the end of follow-up (30 June 2025) were censored at the date of last contact. Patients were divided into a shorter– and a longer-survival group at the Kaplan-Meier median OS of the cohort (< versus ≥ median). The two censored patients had the longest observed follow-up and were therefore unambiguously assigned to the longer-survival group. Analyses were performed on this internal dataset without imputation of missing values. A de-identified version, in which the original dates, the pathology number and other directly identifying variables are removed and age is grouped into 10-year bands, is available upon reasonable request (see Data availability).

### Bioinformatic processing of sequencing data

Amplicon sequencing of the bacterial ribosomal 16S rRNA gene and the fungal internal transcribed spacer (ITS) region was performed by LGC Genomics (Berlin, Germany) using the primer pairs 341F-785R (16S) and fITS7-ITS4R (ITS) on an Illumina MiSeq platform (v3 chemistry, 300 bp paired-end reads). Raw, adapter-trimmed and merged reads were provided by the sequencing company together with per-sample FastQC reports, which were aggregated using MultiQC.^55^ The 16S and ITS data were processed separately with an adapted version of the metabarcoding pipeline,^56,57^ which performs quality filtering, dereplication, denoising, amplicon sequence variant (ASV) generation, chimera removal and taxonomic classification with vsearch (v2.27.0).^58^ Fungal ASVs were classified against UNITE (2022-10-27)^59^ and RDP ITS (v2)^60^, and bacterial ASVs against RDP 16S (v18)^60^ and SILVA^61^ (v123).

One extraction and one buffer control were carried through library preparation and sequencing for each marker. Three of the four controls yielded negligible read numbers (948 and 265 reads for 16S, 166 reads for the ITS extraction control), while the ITS buffer control yielded 4,749 reads, dominated by *Debaryomyces hansenii* and *Heterobasidion abietinum*. Controls were excluded from all analyses. No computational decontamination was applied, as prevalence-based approaches have very limited power with a single control of each type per marker.

### Microbiome analysis

All downstream analyses were conducted in R (4.4.1)^62^ using phyloseq (1.48.0)^63^ and tidyverse (2.0.0).^64^ Samples with fewer than 1,000 reads were excluded. No rarefaction and no abundance– or prevalence-based filtering were applied; only taxa with zero counts across all samples were removed. For community-level analyses, ASVs were agglomerated to genus level (16S) and species level (ITS) after resolving accession-based rank placeholders in the reference taxonomy, unclassified ASVs were retained and labelled at their most resolved available rank (e.g. “Lachnospiraceae (unclassified)”). Such unresolved assignments accounted for 9.1% of counts at genus level (16S) and 6.9% at species level (ITS).

All microbiome analyses were performed separately for both markers (16S and ITS) and for two contrasts: patients versus controls, and, within the patient cohort, patients above versus below the Kaplan-Meier median OS. Alpha diversity (Shannon and Simpson indices) was estimated for each sample with DivNet (0.4.1),^65^ which accounts for taxon co-occurrence and returns a diversity estimate with its standard error per sample. DivNet was fitted with a per-sample (identity) design matrix and the “careful” tuning setting. *Saccharomyces cerevisiae* was specified as the base taxon for ITS, and the default base taxon was used for 16S. Between-group differences were tested with betta (betta_lincom; breakaway v4.8.5) using the contrast as the only fixed effect, which models these estimates together with their standard errors and accounts for heterogeneity between samples^66^. Group means with 95% confidence intervals were derived from the same model. Beta diversity was quantified as DivNet-derived Bray-Curtis dissimilarity (per-specimen design matrix without variance estimation), tested between groups by a bootstrap procedure (testBetaDiversity, 10,000 iterations), and ordinated by principal coordinates analysis (PCoA) with Cailliez correction for negative eigenvalues (ape).^67^ Differentially abundant taxa were identified with the robust score test implemented in radEmu (2.2.1.0).^68^ Given the limited sample size, models were fitted without covariates. As this study was explicitly exploratory, we did not correct for multiple testing and report taxa at a nominal, uncorrected threshold of p ≤ 0.05, interpreting them as candidate signals.

### Statistical analysis of cytokine concentrations and survival analysis

Peripheral cytokine concentrations were compared between patients and controls, and between survival groups using two-sided Wilcoxon rank-sum tests and evaluated as an exploratory panel without correction for multiple testing. To identify tissue markers associated with outcome, we analyzed OS by the Kaplan-Meier method and Cox proportional-hazards regression (survival, 3.8-3). Age at diagnosis, KPS and extent of resection (reference: total resection) were specified as *a priori* confounders. All remaining candidate variables (Iba1, TMEM119, P2RY12, Ki-67 and the percentage of MGMT promoter methylation) were screened in univariate Cox models against a null model without covariates refitted on the same complete cases. A variable was carried forward if the likelihood-ratio test yielded p ≤ 0.05 and the Akaike information criterion decreased by at least two units. Variables showing non-convergence or quasi-separation were discarded. The primary multivariable model included Iba1 immunoreactivity, the only marker retained in the screening, together with the confounders. Continuous predictors were rescaled so that hazard ratios refer to a 1% increase in Iba1-positive cells, a 10-year increase in age and a 10-point increase in KPS.

For the primary model, we examined the proportional-hazards assumption using scaled Schoenfeld residuals, the functional form of continuous predictors using martingale residuals from the null model, influential observations using dfbeta residuals together with a leave-one-out analysis, and collinearity using variance inflation factors.

As sensitivity analyses, we fitted parsimonious models combining Iba1 with each confounder alone and in pairs, and models adding MGMT promoter methylation status (methylated versus unmethylated) either to Iba1 only or to the primary model. The hazard ratio for Iba1 was consistent across all of these models.

### Use of large language models

We used ChatGPT (OpenAI; GPT-5, GPT-5.1, GPT-5.2) and Claude (Anthropic; Claude Opus 4.8, Opus 5, Opus 5.5) to assist with (i) language editing, (ii) literature and background searches, and (iii) programming support, including looking up and adapting analysis code. We critically reviewed, verified, revised and approved all output generated by large language models and take full responsibility for the integrity and content of the manuscript.

## Code availability

All analysis code and the sample group assignments required to reproduce the microbiome analyses are available at https://github.com/viax23/GBM-Microbiome and archived at Zenodo (DOI: 10.5281/zenodo.23417943).

## Data availability

The raw sequencing data generated in this study have been deposited in the European Nucleotide Archive under accession number PRJEB91343.

Individual-level clinical metadata cannot be made publicly available: owing to the small, single-center cohort, these data cannot be reliably anonymized. Aggregate cohort characteristics are provided in Table 1; de-identified data are available from the corresponding author upon reasonable request.

## Authors contributions

Conceptualization: VN, MH, KS. Methodology: VN, MH, KS, CMM, MA, NG, KH. Investigation: MH, VN, KS, ER. Resources: ML, AFK, RN, AC, VN, MH, CH, RIE, OK. Data curation: MH, KS, MA. Formal analysis: MH, VN, KS, MA, ER. Visualization: MH, KS. Supervision: VN, MH, ML, OK. Project administration: VN, MH. Writing – original draft: MH, VN, KS. Writing – review and editing: MH, VN, KS, AFK, AC, RN, ML, CH, RIE, MA, NG, KH, OK, CMM, ER. All authors read and approved the final manuscript.

## Acknowledgements

We are very grateful to Dagmar Hemmerich (Division Experimental Neurosurgery, Department of Neurosurgery, University Hospital Würzburg, Josef-Schneider-Straße 11, 97080 Würzburg, Germany) for excellent technical assistance.

This work was supported by the Interdisciplinary Center for Clinical Research (IZKF), University of Würzburg, under Grant B-450, and by the Bavarian Center for Cancer Research (BZKF). Open Access funding was supported by the Open Access Publishing Fund of the University of Heidelberg. The funders had no role in the study design, data collection, analysis and interpretation, or in writing the manuscript.

## Supplementary Information

Supplementary Information: Supplementary Figures 1–3 and Tables 1–2.

Supplementary Data 1: Differential abundance results. Log-fold changes, 95% confidence intervals, and unadjusted and Benjamini-Hochberg-adjusted p-values from radEmu robust score tests for all tested bacterial genera and fungal species, for patients versus controls and longer versus shorter overall survival.

---

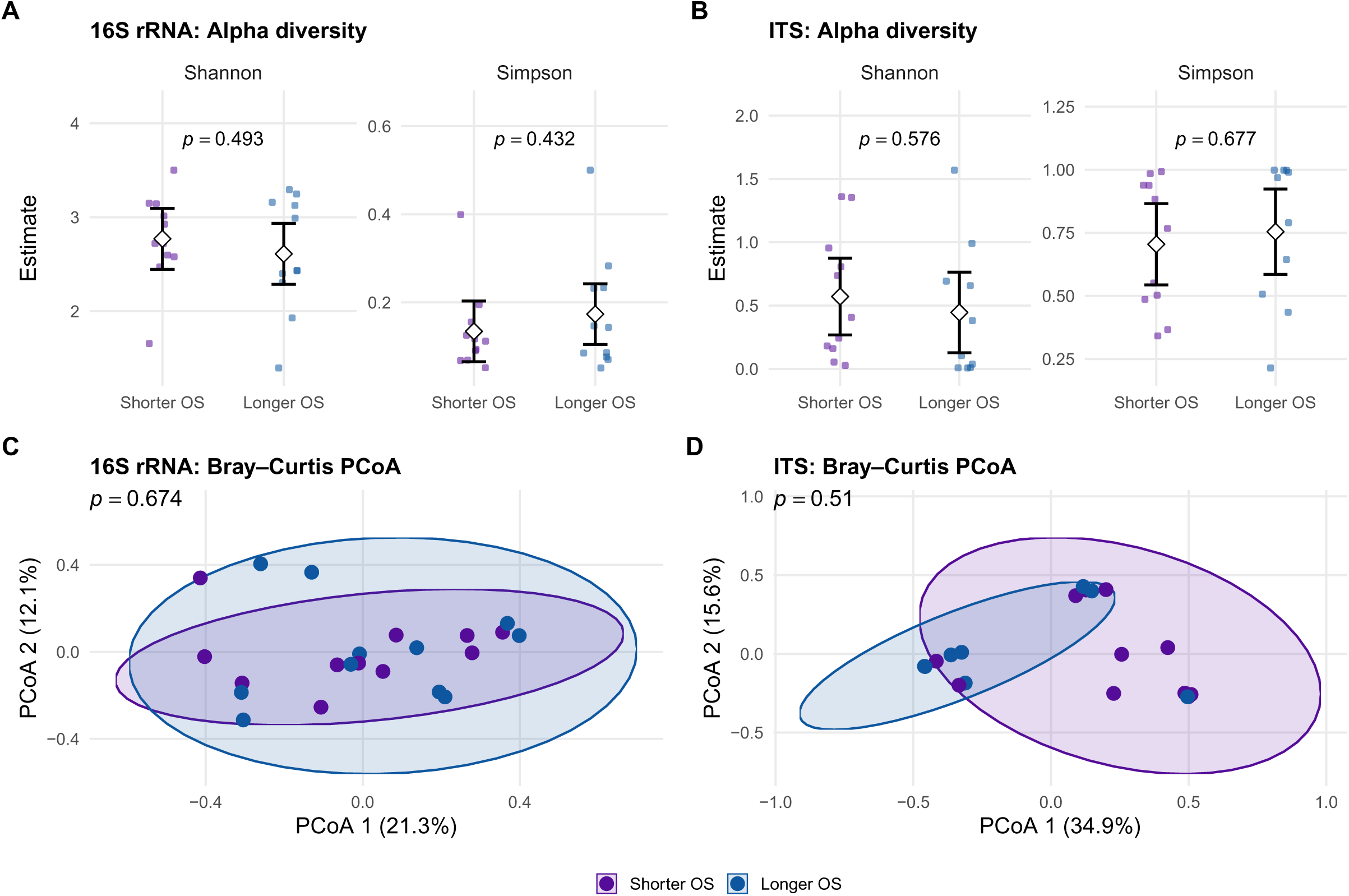

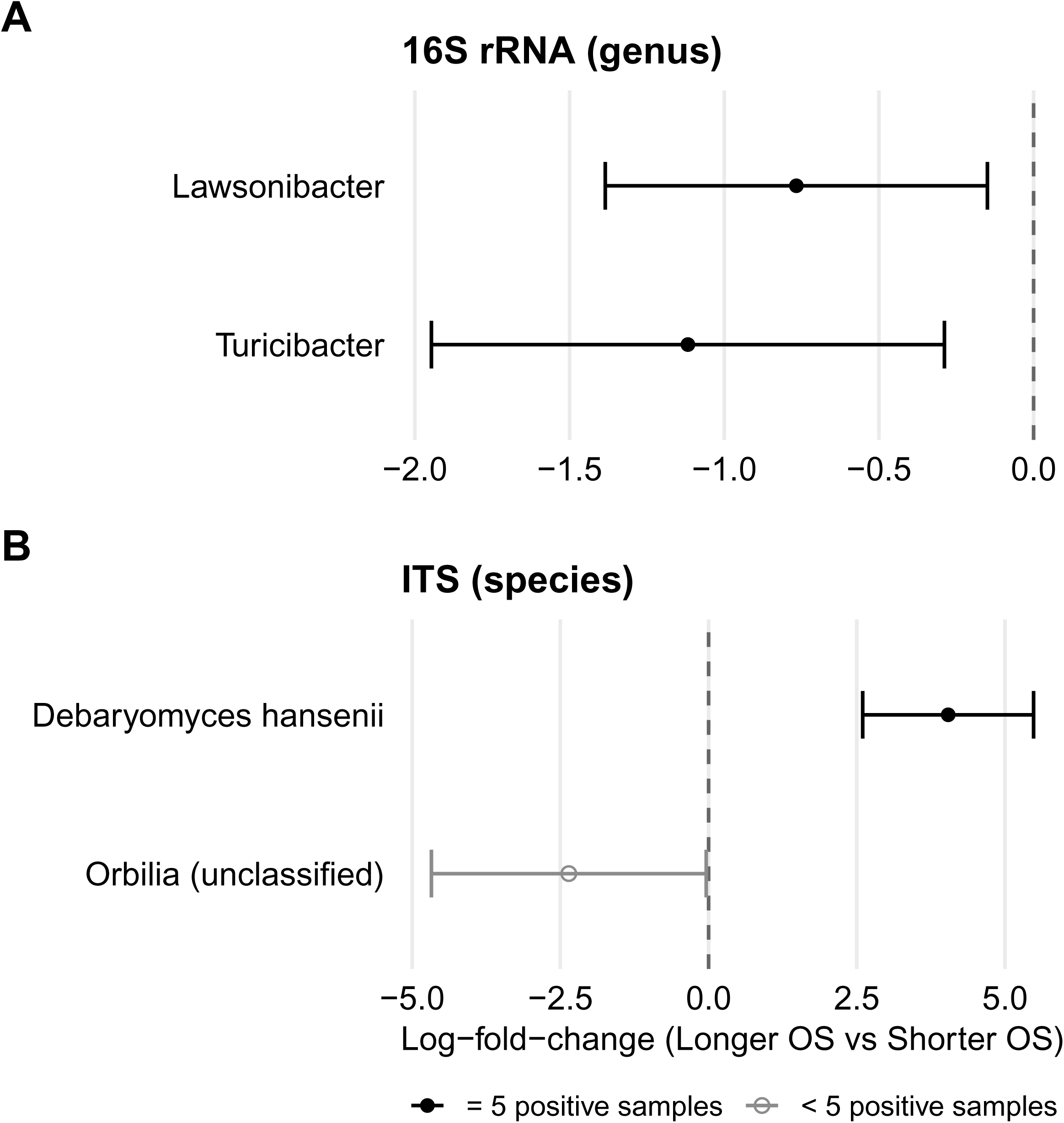

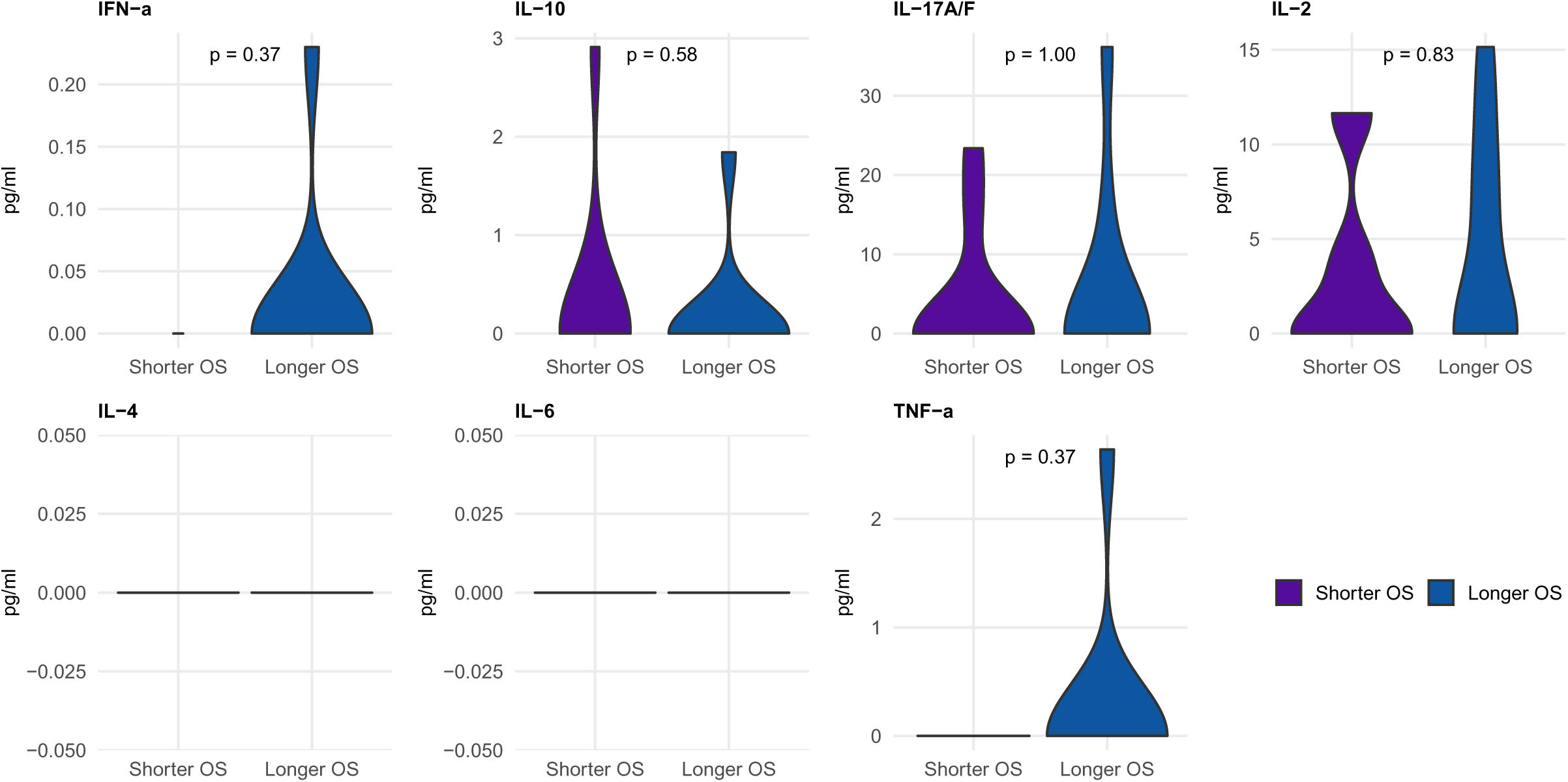

