## Supplementary Information for "A prospective case-control study of the gut bacteriome and mycobiome in glioblastoma"

### Supplementary Figures

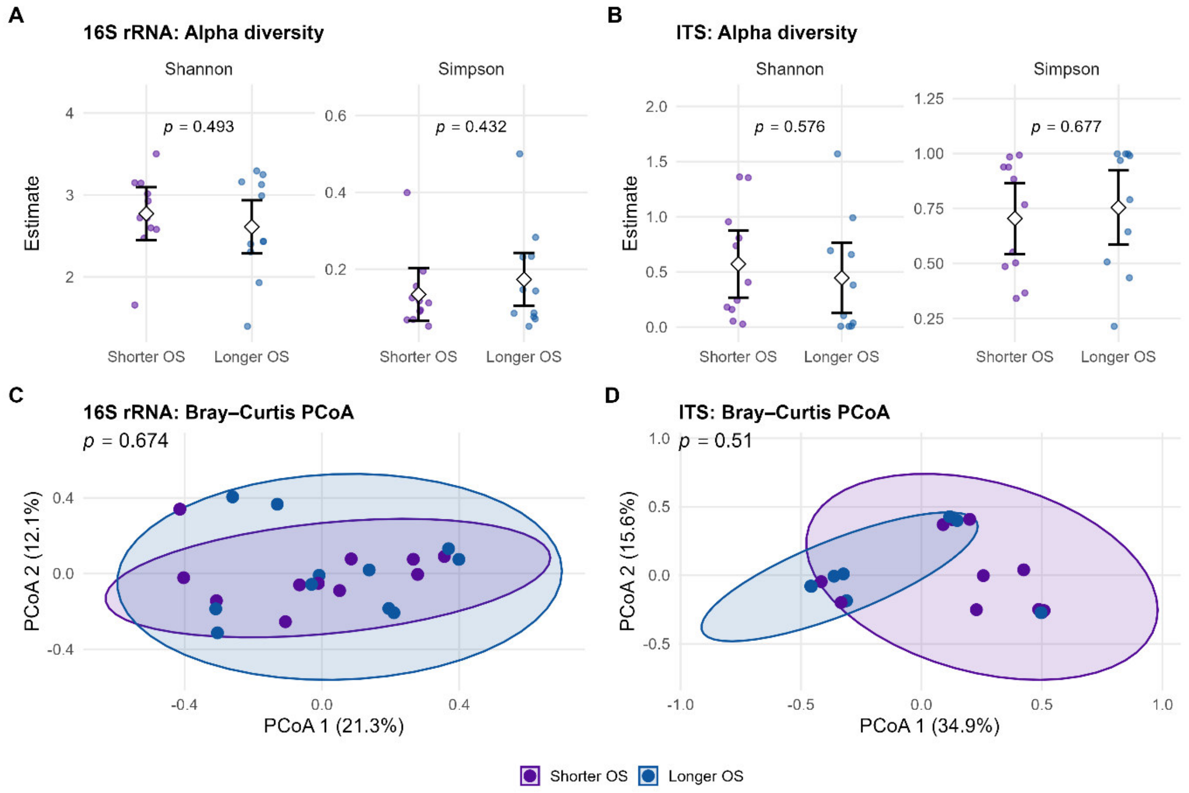


**Supplementary Figure 1. Gut microbiome diversity in patients with shorter and longer OS.**

**(A, B)** Shannon and Simpson diversity for (A) 16S rRNA and (B) ITS; small points show per-sample DivNet estimates, diamonds and error bars the group means with 95% confidence intervals estimated by betta; p-values from betta. **(C, D)** Principal coordinates analysis of Bray-Curtis dissimilarities for (C) 16S rRNA and (D) ITS; ellipses indicate 95% confidence regions. Survival groups were split at the cohort's median OS.

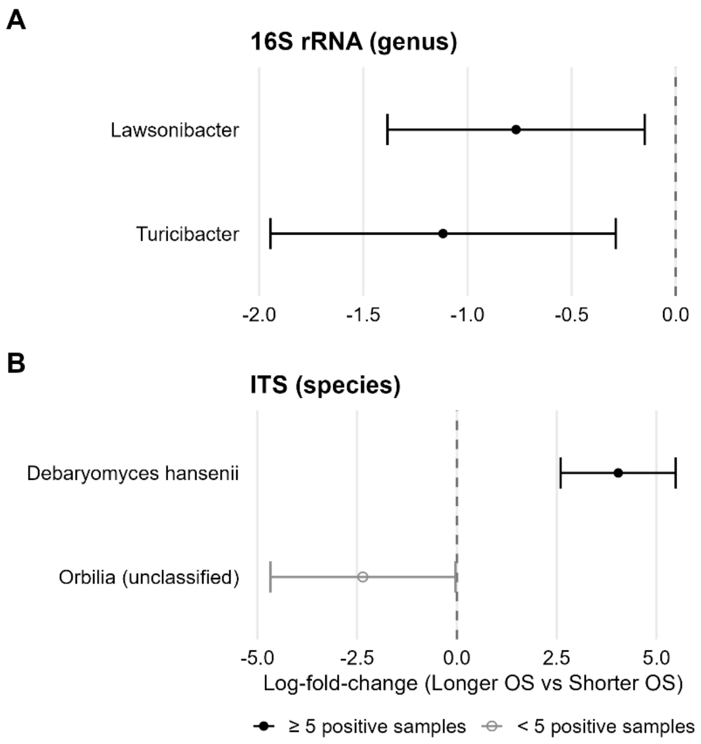


**Supplementary Figure 2. Taxa differing in abundance between patients with shorter and longer OS.**

Log-fold-changes with 95% confidence intervals for (A) bacterial genera and (B) fungal species; positive values indicate higher abundance in the longer-survival group. All taxa with an uncorrected $p \leq 0.05$ are shown; open symbols mark taxa detected in fewer than five samples.

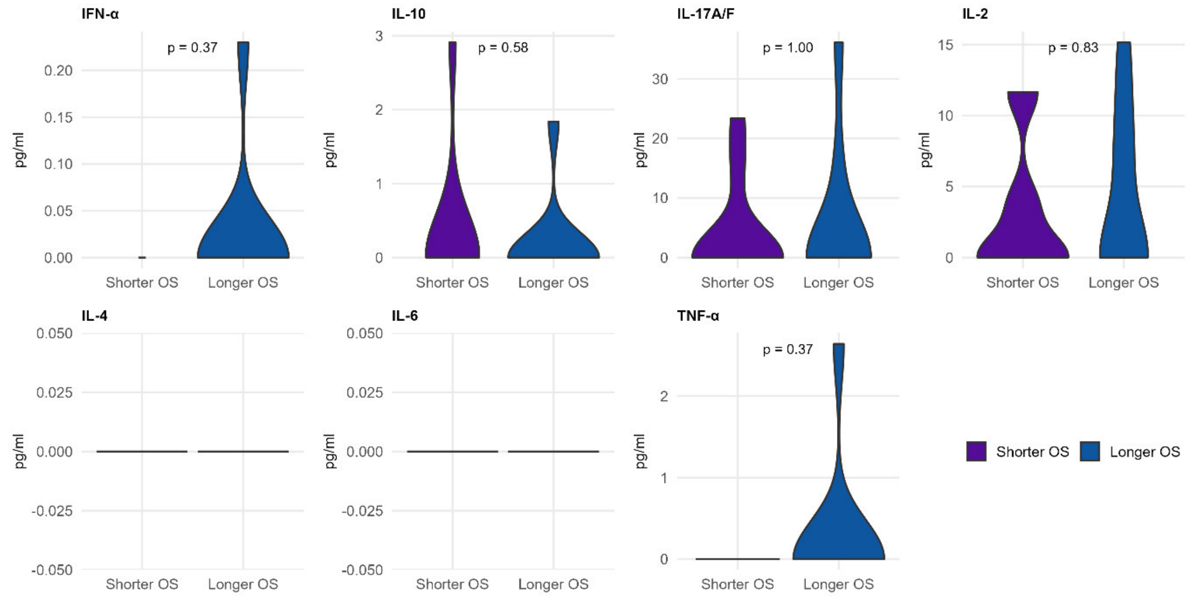


**Supplementary Figure 3. Plasma cytokine concentrations by survival group.**

Concentrations of seven cytokines in patients with shorter and longer OS; p-values from Wilcoxon rank-sum tests.

### Supplementary Tables

| **Variable** | **n** | **Events** | **HR (95% CI)** | **LRT p** | **Δ AIC** |
|---|---|---|---|---|---|
| Iba1 (per 1% of cells) | 21 | 19 | 0.83 (0.72-0.97) | 0.02 | -3.49 |
| TMEM119 (per 1% of cells) | 21 | 19 | 0.99 (0.93-1.06) | 0.87 | 1.97 |
| P2RY12 (per 1% of cells) | 21 | 19 | 0.98 (0.90-1.06) | 0.53 | 1.61 |
| Ki-67 (per 1% of cells) | 23 | 21 | 0.96 (0.88-1.05) | 0.37 | 1.18 |
| MGMT promoter methylation (per 1 %) | 5 | 5 | 1.06 (0.98-1.13) | 0.06 | -1.63 |

**Supplementary Table 1. Screening of candidate immunohistochemical and molecular markers for association with OS.**

Shown are the number of patients with available data (n), the number of deaths (events), the hazard ratio (HR) with 95% confidence interval, the likelihood ratio test (LRT) p-value against the null model, and the difference in Akaike information criterion (AIC) between the screening and the null model (ΔAIC; negative values indicate improved fit). Variables with LRT $p \leq 0.05$ and $\Delta AIC \leq -2$ were carried forward into multivariable modelling. AIC, Akaike information criterion; CI, confidence interval; HR, hazard ratio; Iba1, ionized calcium-binding adapter molecule 1; Ki-67, proliferation marker Ki-67; LRT, likelihood ratio test; MGMT, O-6-methylguanine-DNA methyltransferase; OS, overall survival; P2RY12, purinergic receptor P2Y12; TMEM119, transmembrane protein 119.

| Model | Iba1 (per 1% of cells) HR (95% CI) | Iba1 p-value |
|---|---|---|
| Primary (Iba1 + Resection + Age + KPS) | 0.82 (0.67-0.99) | 0.04 |
| Iba1 + Age | 0.81 (0.69-0.95) | 0.01 |
| Iba1 + KPS | 0.84 (0.72-0.98) | 0.03 |
| Iba1 + Resection | 0.87 (0.73-1.03) | 0.11 |
| Iba1 + Age + Resection | 0.82 (0.68-0.99) | 0.04 |
| Iba1 + KPS + Resection | 0.88 (0.73-1.05) | 0.15 |
| Iba1 + Age + KPS | 0.81 (0.69-0.95) | 0.01 |
| Iba1 + Methylation | 0.83 (0.72-0.97) | 0.02 |
| Iba1 + Age + KPS + Resection + Methylation | 0.82 (0.67-1.02) | 0.07 |

**Supplementary Table 2.** Sensitivity analysis of the association between Iba1 and OS.

Shown are hazard ratios (HR) with 95% confidence intervals and p-values for Iba1 (per 1% increase in Iba1-positive cells) from Cox proportional hazards models with varying covariate sets. The primary model was adjusted for extent of resection, age at diagnosis, and Karnofsky performance status (KPS). Methylation refers to MGMT promoter methylation status. P-values are unadjusted for multiple comparisons, as all models test the same association and serve a sensitivity rather than a confirmatory purpose. CI, confidence interval; HR, hazard ratio; Iba1, ionized calcium-binding adapter molecule 1; KPS, Karnofsky performance status; MGMT, O-6-methylguanine-DNA methyltransferase; OS, overall survival.
